# Applying Multimodal Data Fusion based on Deep Learning Methods for the Diagnoses of Neglected Tropical Diseases: A Systematic Review

**DOI:** 10.1101/2024.01.07.24300957

**Authors:** Yohannes Minyilu, Mohammed Abebe, Million Meshesha

## Abstract

Neglected tropical diseases (NTDs) are the most prevalent diseases worldwide affecting one-tenth of the world population. Although there are multiple approaches to diagnosing these diseases, using skin manifestations and lesions caused as a result of these diseases along with other medical records is the preferred method. This fact triggers the need to explore and implement a deep learning-based diagnostic model using multimodal data fusion (MMDF) techniques to enhance the diagnostic process. This paper, thus, endeavored to present a thorough systematic review of studies regarding the implementation of MMDF techniques for the diagnoses of skin-related NTDs. To achieve its objective, the study used the PRISMA method based on predefined questions and collected 427 articles from seven major and reputed sources and critically appraised each article. Since no previous studies were found regarding the implementation of MMDF for the diagnoses of skin related NTDs, similar studies using MMDF for the diagnoses of other skin diseases, such as skin cancer, were collected and analyzed in this review to extract information about the implementation of these methods. In doing so, various studies are analyzed using six different parameters including research approaches, disease selected for diagnosis, dataset, algorithms, performance achievements and future directions. Accordingly, although all the studies used diverse research methods and datasets based on their problem, deep learning-based convolutional neural networks (CNN) algorithms are found to be the most frequently used and best performing models in all studies reviewed.

## 1. Introduction

Being the largest organ in the human body, the skin can serve as an indicator of some illnesses arising from different causes such as cancer, internal organ failure, and neglected tropical diseases. NTDs are the most prevalent diseases globally affecting more than one billion people worldwide (i.e., more than ten percent of the world’s population), particularly, in the tropical areas of the world among the poorest, most vulnerable and outcast groups and still have devastating impacts on people’s physical, mental, and social well-being [1][2][3][4]. However, these diseases can be diagnosed using skin related symptoms since majority of the NTDs have primary skin indicators or associated clinical features where 18 of the 20 NTDs (recognized by the World Health Organization (WHO)) having skin related symptoms [5]. Hence, the utilization of DL-based diagnostic systems for the diagnoses and recognition of skin related NTDs will be a great achievement in overcoming the NTDs. This study endeavored to present a thorough systematic review of studies regarding the implementation of MMDF techniques for the diagnoses of skin related NTDs. Since no previous studies implemented MMDF techniques for the diagnoses of skin NTDs, related studies conducted for the diagnoses of skin diseases other than NTDs using MMDF and DL methods were deeply appraised by this review. These studies confirmed that the utilization of MMDF techniques outperforms the traditional diagnostic models that implemented DL methods without MMDF [6][7][8][9]. It is in view of these facts the study is -motivated to conduct this systematic literature review and a thorough appraisal of previous studies using the PRISMA method of systematic review based on the following guiding questions:

- What DL methods or approaches were utilized for the diagnoses of the skin disease(s)?
- Which data fusion methods were used for the skin disease diagnosis tasks?
- What types of medical data were integrated to demonstrate MMDF method for the diagnoses of the skin diseases?
- Which algorithms were used and how does each algorithm perform in the DL-based MMDF skin disease diagnostic model or system?

## 2. The Need for Intelligent Diagnostic Systems

In recent times, due to the high desire to enhance the diagnostic processes in the healthcare sectors, the utilization of automated and intelligent diagnostic systems is getting greater attention for the diagnosis of various diseases. In this regard, intelligent diagnostic systems built based on machine learning (ML) and deep learning (DL) methods are the most researched and deployed approaches in the healthcare sector to support diagnostic decision making. On the other hand, in the real-world clinical settings, efficient disease diagnostic processes are basically carried out by using different clinical data that are taken from different sources and different formats or modalities including textual patient information and medical clinical images such as X-ray, dermoscopic images or even patient skin images. The integrative utilization of the diverse modalities of medical data can be used to enhance the diagnostic processes, thereby enhancing the quality of healthcare services, by using the ML and DL methods. In ML, this process of integrating multiple modalities of data (possibly taken from different sources) is technically called multimodal data fusion [10][11]. Multimodal data techniques are playing vital roles in developing intelligent disease diagnostic systems for different diseases such as in dermatology [12]. In this regard, MMDF techniques are advancing diagnostic accuracy, where these methods outperform other baseline methods, as presented in [13].

### 2.1. Deep Learning and Diagnoses of NTDs

The current diagnostic approaches used for NTDs are mainly based on clinical procedures, such as patient observation and laboratory examinations based on limited resources in most affected areas. Currently, however, there are efforts towards the utilization of intelligent diagnostic tools using ML and DL approaches. Since most of the NTDs are curtly being diagnosed using skin manifestation, the utilization of DL-based approaches for the diagnoses of these diseases would be a great potential to support and enhance the diagnostic processes. In this regard, different studies were previously conducted to diagnose various NTDs. Beesetty et al. [14], conducted a study towards leprosy skin lesion detection employing a Siamese (Siamese NN)-based few-shot learning (FSL) model for a small clinical dataset and claimed a higher (91.25%) diagnostic accuracy. On the other hand, Ali et al. [15] used ML methods for early prediction of Schistosomiasis and concluded with the CatBoost model showing the best performance with the highest accuracy of 87.1%. An optimized diagnostic approach was also proposed for NTDs by selecting three diseases and developing a model using SVM and the black hole algorithm (BHO) achieving an accuracy of 96% [16]. Another study reviewed by the current study demonstrated a DL-based diagnostic model for NTDs using skin images only and achieved 70% accuracy [17]. All the aforementioned studies utilized ML and DL methods for the diagnoses of NTDs and achieved remarkable results in terms of accuracy. However, no previous studies were found that utilized DL-based methods using MMDF techniques which will help to achieve higher diagnostic accuracies, as experimented in other studies for non-NTD skin diseases which require further research.

### 2.2. Data Fusion Approaches

Data or information fusion represents the usage of data or information from different sources in different formats or modalities for interpretation in all tasks that require any type of parameter estimation or prediction using data or information [18]. There are different fusion techniques to combine and aggregate multimodal data which include feature-level fusion, decision-level or late fusion, hybrid multimodal fusion, model-level fusion, rule-based fusion, classification-based fusion and estimation-based fusion [19].

#### 2.2.1. Feature Fusion

Feature fusion is a data integration technique used to aggregate multiple feature sets extracted from multiple input data to generate a single feature set [19]. In image processing problems, it refers to the fusion of feature vectors of training images extracted from shared weight network layer and feature vectors composed of other numerical data [20]. It helps to learn image features fully for the description of their rich internal information [21]. Various studies are found and appraised that use feature fusion techniques to develop diagnostic models for the diagnoses of skin diseases, as summarized in Table 1.

**Table 1:** Review of the future fusion and related techniques for skin disease diagnoses.

| Ref | Pub. Yr. | Study Method / Approach Used | Disease(s) Selected | Dataset(s) Used | Algorithm(s) Used | Performance Results Achieved |
| --- | --- | --- | --- | --- | --- | --- |
| [31] | 2019 | Transfer Learning and multi-layer **feature fusion** network | Skin Lesion | HAM1000<br>0 dataset | CNN | high recognition (ROC-AUC 96.51) |
| [24] | 2021 | **Image fusion (clinical & dermoscopic)**: multi-labeled deep feature extractor and clinically constrained classifier chain (CC) | Skin Cancer (Melanoma) | publicly available 7-point checklist dataset | DCNN, CC, PCA | Reported 81.3% accuracy |
| [6] | 2022 | Multiclass skin lesion classification using **feature fusion** & extreme learning machine (ELM) | Skin Disease (Skin Lesion) | HAM1000<br>0 and ISIC2018 | SVM, fine KNN, DT, NB, ensemble tree (EBT), & single hidden layer ELM | Registered best accuracy of 94.36 percent |
| [32] | 2022 | **Apply features fusion on** manual and automatic feature extraction | Skin Cancer | DermIS dataset | CNN, LSTM, LBP, LBP, Inception V3 | Achieved maximum accuracy of 99.4% |
| [33] | 2023 | **Dual-branch (feature fusion)** network using DCNN and Transformer branches for local and global feature extraction | Skin Disease (Skin Lesion) | Used a private dataset XJUSL | DCNN | Reducing the number of parameters by 11.17 M improved classification accuracy by 1.08% |
| [34] | 2023 | **Feature fusion**: fast-bounding box (FBB), Hybrid Feature Extractor (HFE), and the CNN VGG19 based CNN | Skin Cancer (Melanoma) | ISIC 2017 and the Academic torrents dataset | CNN | Registered 99.85% accuracy |

#### 2.2.2. Model Fusion

Model fusion, also known as late fusion, represents a fusion approach that combines different models. The study done by AlDahoul et al. [22], combines two deep neural networks including binary normal/attack classifier and multi-attack classifier to train a deep neural network (DNN) for network anomaly detection. As mentioned in [19], the model fusion technique uses the connection between experimental data under different modalities.

#### 2.2.3. Image Fusion

Image fusion combines different images and generates informative images by integrating images obtained from different sources [23]. A previous study [24], suggested that aggregating medical images helps to enhance diagnostic accuracy. This claim was demonstrated by fusing clinical images and dermoscopic images using the deep convolutional neural networks (DCNN) methods and achieved an overall accuracy of 81.3%. While the clinical images are clinically captured photographs [25], dermoscopic images represent images taken by dermatologists using dermoscopy [26].

#### 2.2.4. Multimodal Data Fusion

Multimodal data represents the different formats or modalities of data such as text, image, video, and audio. A multimodal data fusion approach is used for combining particular modalities to derive multimodal representation [10][11][19][27]. This approach has multiple applications for healthcare systems as it allows the combination of different modalities of data, such as textual medical history of patients, clinical images of patients (such as skin images of patients) to form a single multimodal dataset that can be used to train diagnostic models using ML and DL methods. In this regard, various studies implemented and demonstrated MMDF for the diagnoses of different skin diseases as summarized in Table 2.

**Table 2:**
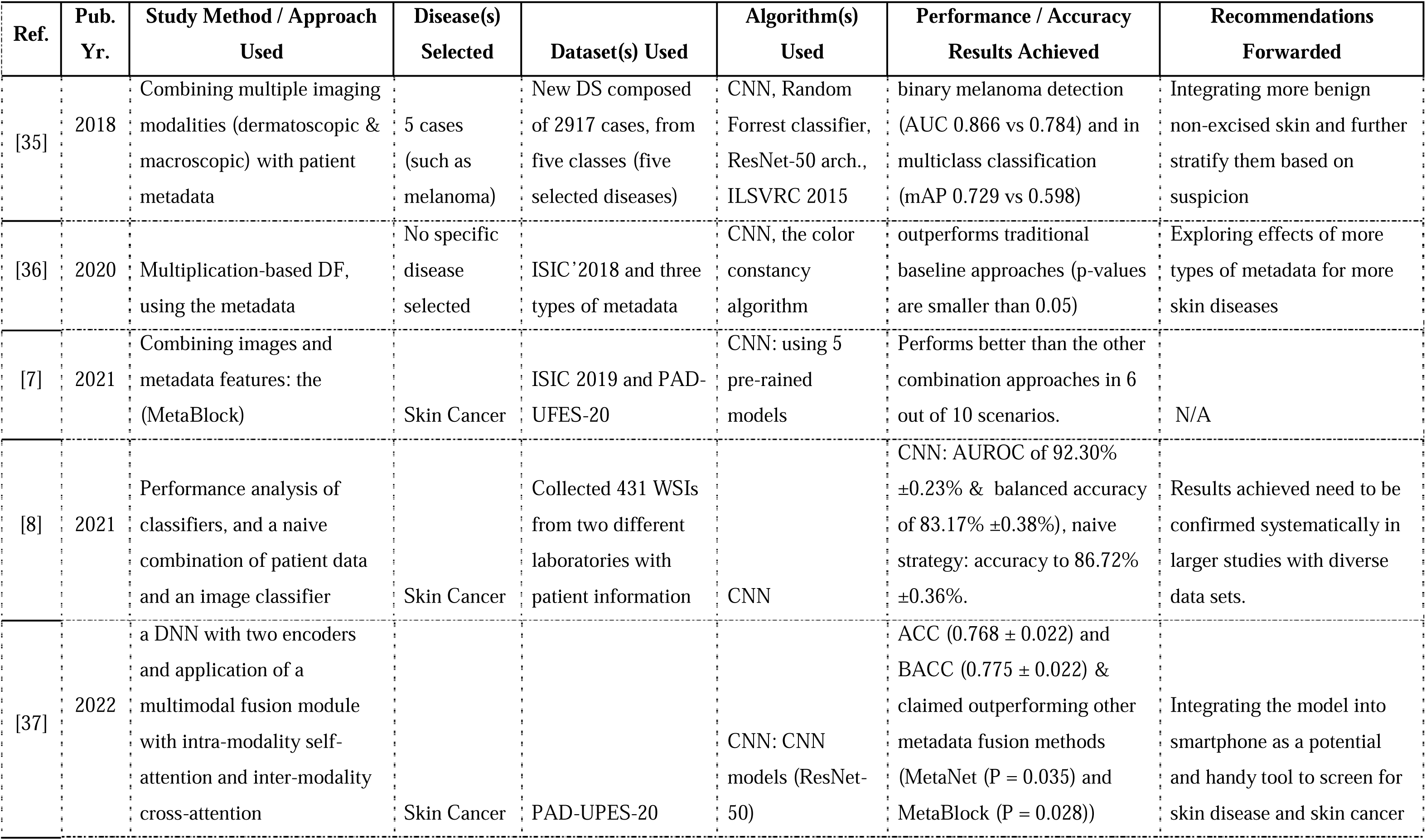
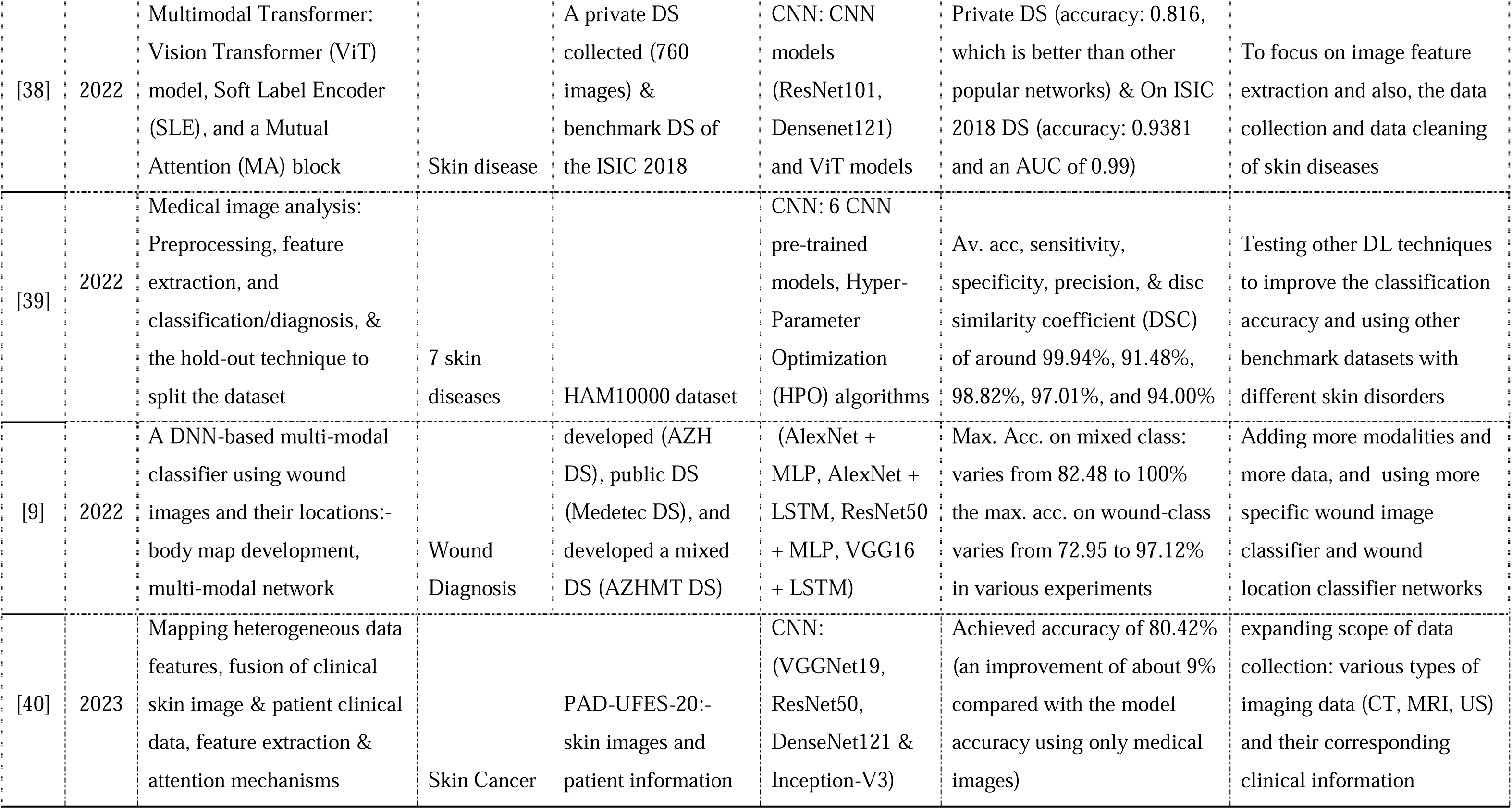
Review of the DL-based multimodal data fusion techniques for the diagnosis of skin diseases.

| Ref. | Pub. Yr. | Study Method / Approach Used | Disease(s) Selected | Dataset(s) Used | Algorithm(s) Used | Performance / Accuracy Results Achieved | Recommendations Forwarded |
| --- | --- | --- | --- | --- | --- | --- | --- |
| [35] | 2018 | Combining multiple imaging modalities (dermatoscopic & macroscopic) with patient metadata | 5 cases (such as melanoma) | New DS composed of 2917 cases, from five classes (five selected diseases) | CNN, Random Forrest classifier, ResNet-50 arch., ILSVRC 2015 | binary melanoma detection (AUC 0.866 vs 0.784) and in multiclass classification (mAP 0.729 vs 0.598) | Integrating more benign non-excised skin and further stratify them based on suspicion |
| [36] | 2020 | Multiplication-based DF, using the metadata | No specific disease selected | ISIC'2018 and three types of metadata | CNN, the color constancy algorithm | outperforms traditional baseline approaches (p-values are smaller than 0.05) | Exploring effects of more types of metadata for more skin diseases |
| [7] | 2021 | Combining images and metadata features: the (MetaBlock) | Skin Cancer | ISIC 2019 and PAD-UFES-20 | CNN: using 5 pre-trained models | Performs better than the other combination approaches in 6 out of 10 scenarios. | N/A |
| [8] | 2021 | Performance analysis of classifiers, and a naive combination of patient data and an image classifier | Skin Cancer | Collected 431 WSIs from two different laboratories with patient information | CNN | CNN: AUROC of 92.30% $\pm 0.23\%$ & balanced accuracy of 83.17% $\pm 0.38\%$, naive strategy: accuracy to 86.72% $\pm 0.36\%$. | Results achieved need to be confirmed systematically in larger studies with diverse data sets. |
| [37] | 2022 | a DNN with two encoders and application of a multimodal fusion module with intra-modality self-attention and inter-modality cross-attention | Skin Cancer | PAD-UPES-20 | CNN: CNN models (ResNet-50) | ACC (0.768 ± 0.022) and BACC (0.775 ± 0.022) & claimed outperforming other metadata fusion methods (MetaNet (P = 0.035) and MetaBlock (P = 0.028)) | Integrating the model into smartphone as a potential and handy tool to screen for skin disease and skin cancer |

|  |  |  |  |  |  |  |  |
| --- | --- | --- | --- | --- | --- | --- | --- |
| [38] | 2022 | Multimodal Transformer: Vision Transformer (ViT) model, Soft Label Encoder (SLE), and a Mutual Attention (MA) block | Skin disease | A private DS collected (760 images) & benchmark DS of the ISIC 2018 | CNN: CNN models (ResNet101, Densenet121) and ViT models | Private DS (accuracy: 0.816, which is better than other popular networks) & On ISIC 2018 DS (accuracy: 0.9381 and an AUC of 0.99) | To focus on image feature extraction and also, the data collection and data cleaning of skin diseases |
| [39] | 2022 | Medical image analysis: Preprocessing, feature extraction, and classification/diagnosis, & the hold-out technique to split the dataset | 7 skin diseases | HAM10000 dataset | CNN: 6 CNN pre-trained models, Hyper-Parameter Optimization (HPO) algorithms | Av. acc, sensitivity, specificity, precision, & disc similarity coefficient (DSC) of around 99.94%, 91.48%, 98.82%, 97.01%, and 94.00% | Testing other DL techniques to improve the classification accuracy and using other benchmark datasets with different skin disorders |
| [9] | 2022 | A DNN-based multi-modal classifier using wound images and their locations:- body map development, multi-modal network | Wound Diagnosis | developed (AZH DS), public DS (Medetec DS), and developed a mixed DS (AZHMT DS) | (AlexNet + MLP, AlexNet + LSTM, ResNet50 + MLP, VGG16 + LSTM) | Max. Acc. on mixed class: varies from 82.48 to 100% the max. acc. on wound-class varies from 72.95 to 97.12% in various experiments | Adding more modalities and more data, and using more specific wound image classifier and wound location classifier networks |
| [40] | 2023 | Mapping heterogeneous data features, fusion of clinical skin image & patient clinical data, feature extraction & attention mechanisms | Skin Cancer | PAD-UFES-20:- skin images and patient information | CNN: (VGGNet19, ResNet50, DenseNet121 & Inception-V3) | Achieved accuracy of 80.42% (an improvement of about 9% compared with the model accuracy using only medical images) | expanding scope of data collection: various types of imaging data (CT, MRI, US) and their corresponding clinical information |

## 3. Materials and Methods

To conduct and report this systematic review, we follow as a basis, the steps suggested by the Preferred Reporting Items for Systematic Reviews and Meta-Analyses (PRISMA) model [28][29] as described below.

### 3.1. Search Strategy

Research articles were searched from all the major indexing databases and search engines, such as Google Scholar, PubMed, IEEE Explore, and ScienceDirect. In addition, proper search keywords were prepared and used while searching for the articles, where the search keywords have the appropriate level of relationships with the topics and contents of the articles. A set of searching keywords have been used to deeply search and filter the articles. In this regard, Boolean operators “AND” and “OR” were mainly used. The “AND” operator was used to search for articles in a specific research area to narrow down the search results; “multimodal medical data” AND “data fusion” to search for articles containing both phrases. On the other hand, the “OR” Boolean operator was used to search for articles from wider perspectives as this operator broadens the search results such as “machine learning” OR “deep learning”.

Using the specified methods and operators, multiple search keywords were initially prepared and used to find a sufficient amount of relevant articles. Although a lot of search keywords were used while searching for the articles, some of the keywords include [“Neglected Tropical Diseases” OR “NTDs” AND “Diagnosis” OR “diagnostic model” AND “Deep Learning” OR “DL” OR “Convolutional Neural Network” OR “CNN” OR “Deep Neural Network” OR “DNN” OR “Recurrent Neural Network” OR “RNN”], [”Neglected Tropical Diseases” OR “NTDs” AND “Diagnosis” OR “diagnostic model” AND “Deep Learning” OR “DL” or “Convolutional Neural Network” OR “CNN” OR “Deep Neural Network” OR “DNN” OR “Recurrent Neural Network” OR “RNN” AND “Data Fusion” OR “Multimodal medical Data” OR “Multimodal Data Fusion”], [(((deep learning) AND ((diagnostic model) OR (diagnostic system) OR (diagnostic tool)) AND (skin diseases) AND (skin images)) AND (medical record)) AND (data fusion))].

### 3.2. Eligibility Criteria

Out of the total 427 selected articles, not all articles are critically relevant for the review concerning the integration of multimodal data fusion techniques based on DL methods for the diagnosis of skin related NTDs. Hence, a set of inclusion and/or exclusion criteria are applied, as shown below.

- Articles must use and demonstrate DL methods for the diagnosis of skin NTDs, at least skin diseases if not implemented for skin NTDs, with proper evaluation of the methods used.
- Articles must use and demonstrate proper utilization of multimodal data fusion techniques for the diagnosis of skin NTDs, at least skin diseases since there are no previous studies that use multimodal data fusion techniques for the diagnosis of skin NTDs so far.
- Articles must incorporate precise presentation or discussion and evaluation of all the methods and techniques used in that particular article.
- An Article that used DL methods for the diagnosis of skin related diseases other than NTDs is selected if that particular article uses new or emerging DL methods and presents a proper analysis of the methods and techniques used for the diagnosis of that particular skin disease(s). However, articles that use the popular and previously used DL methods for the diagnosis of diseases other than skin diseases have fewer chances to be selected.
- The article should have an appropriate level of similarity and relationships in its topics and contents with the searching keywords used to deeply search and filter the articles.
- Articles that do not utilize DL and data fusion techniques are excluded from analysis.
- Articles published in languages other than English are excluded from analysis.
- Finally, articles published prior to the year 2014 are also excluded.

### 3.3. Article Search

Different searching methods such as ‘basic search’ and ‘advanced search’ methods were used on multiple article sources. First, the ordinary or basic searching method was used where general titles and the proposed keywords were entered in the regular ‘search box’ of each of the databases and searched. Secondly, the ‘advanced search’ option was used which allows to specify subject areas, related topics, publication dates and other relevant options which helps to obtain articles that are relevant to the topic by narrowing down the search results.

Using both of the search methods and search keywords, a thorough and rigorous searching was conducted on multiple search engines, journals, databases and libraries to find relevant articles. The sources include Google Scholar, IEEE Explore, MDPI, Mendeley, Nature, PubMed, ScienceDirect, AJOL, IDP, NCBI, PLOS, Springer, and Tropical Medicine and Health. Finally, by specifying article publication dates and applying the searching methods on the different databases, 427 articles that were published between the year 2014 and 2024 were collected and prepared for screening. Each database was used independently to search articles. In this regard, Google Scholar was primarily used and it allowed us to collect 178 articles from the aforementioned different sources. Furthermore, extensive searches were done in search of articles that implement multimodal data fusion for the diagnosis of skin NTDs. However, no relevant articles were found related to this particular area.

On the other hand, previously searched sources such as academic web portals, academic libraries and research sites were also used as there were relevant documents from these sources. Hence, 16 articles were collected from such sources.

### 3.4. Relevant Article Selection

To select relevant articles, an extensive searching method was used using wider options of searching keywords. The entire process of article selection for this review was conducted based on the PRISMA method as it was an evidence-based minimum set of items for reporting for systematic reviews and meta-analyses [29][28]. This was done to provide insights for the recent and future research regarding the utilization of DL methods for NTDs diagnosis, the integration of data fusion techniques if they have been used for NTDs and finally to assess and present feedback so as to enhance the performances of such DL-based models.

A series of screening operations were implemented on the collected articles in order to identify the most relevant set of articles for this review. In this case, the first level screening was conducted manually on a total of 427 files using file names and titles of the article. This task allowed us to check if there were duplicates as there were similar files downloaded from multiple sources and it was performed manually by opening and checking files. In this process, 397 items were selected out of the total 427 articles in three phases as summarized in Figure 2 below.

**Fig 1:**
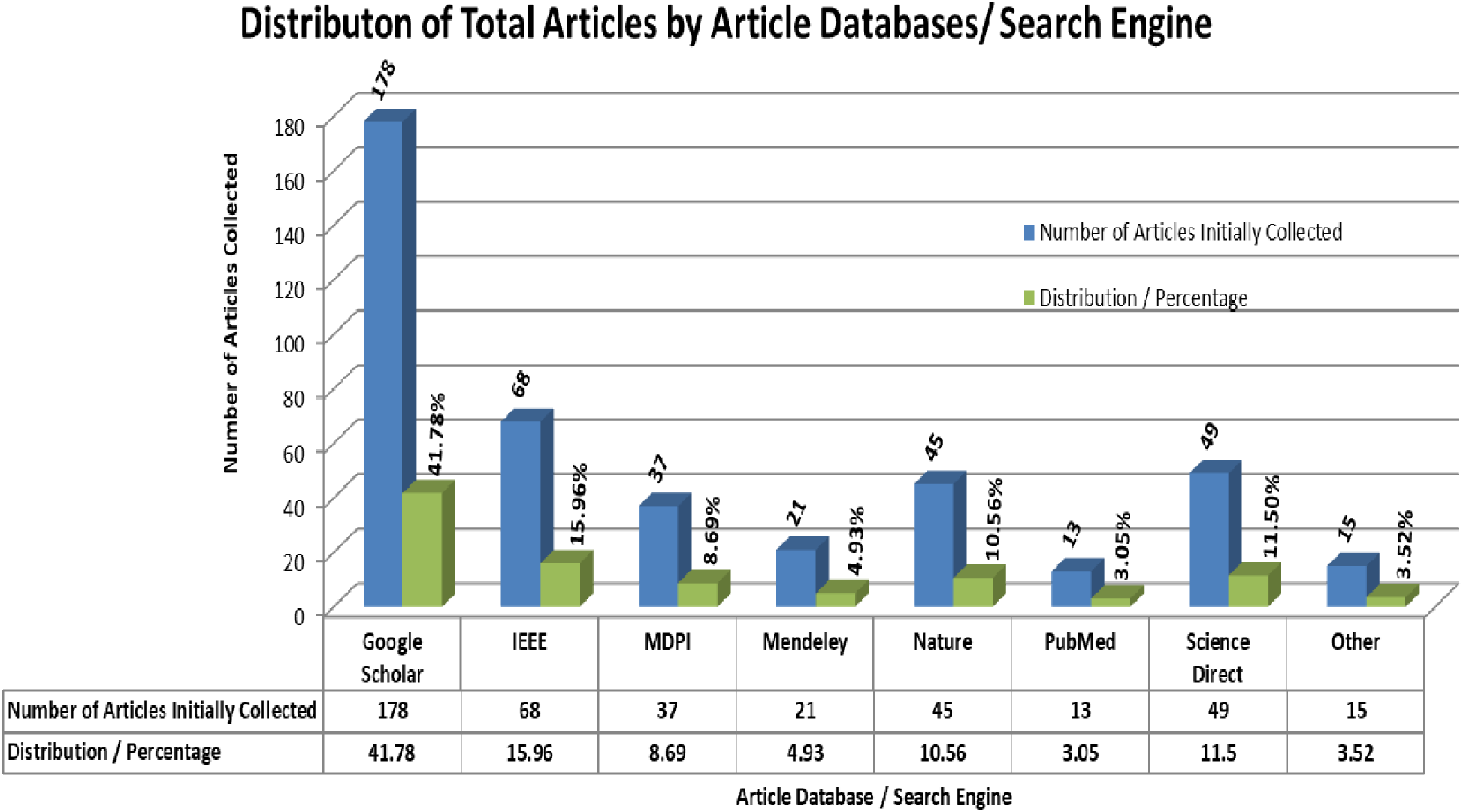
Total collected articles and their distribution by article databases/ search engines.

**Fig 2:**
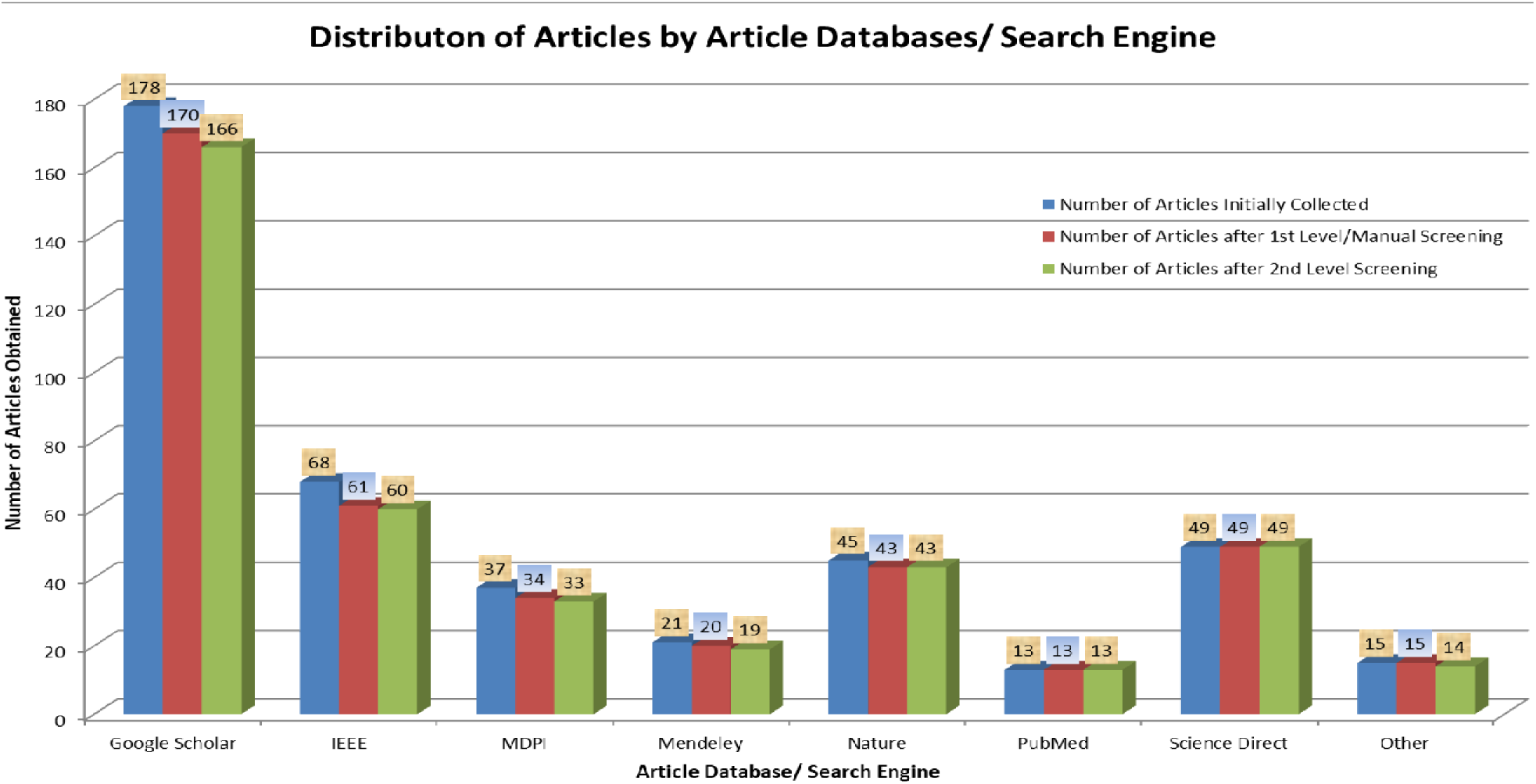
Distribution of articles after first and second level screening.

Then, the next levels of screenings were performed using software tools such as ‘EndNote’ and ‘Rayyan’. As a reference management tool, EndNote was used to create a library containing the collected articles and for manipulation and data processing to check duplicate files in the library. It automatically removed 25 articles as there were duplicate files from different folders followed by an automatic duplicate detection where one duplicate article was identified by EndNote and removed leading to a library containing 371 articles.

The next task involved screening the articles using a higher-level screening software tool based on title, author names and abstracts. For this purpose, Rayyan, a free online software tool [30] was employed which is mainly used to speed up the literature screening process in systematic reviews. This online tool uses the article library exported from EndNote and it was first used to check duplication. Through this process, 4 duplicated articles were detected in the library and two of them were removed where 369 articles were finally identified using Rayyan for the final screening process. Next, using this online software tool, 90 articles that have a relationship with the current topic of the study were selected based on title and abstract analysis. Further screening was required to identify articles in relation to the study area and 18 articles were identified out of the 90 related articles. Finally, 9 articles were selected for the final analysis. The overall article selection procedure is outlined using the PRISMA flow chart as depicted in Figure 3 below.

**Fig 3:**
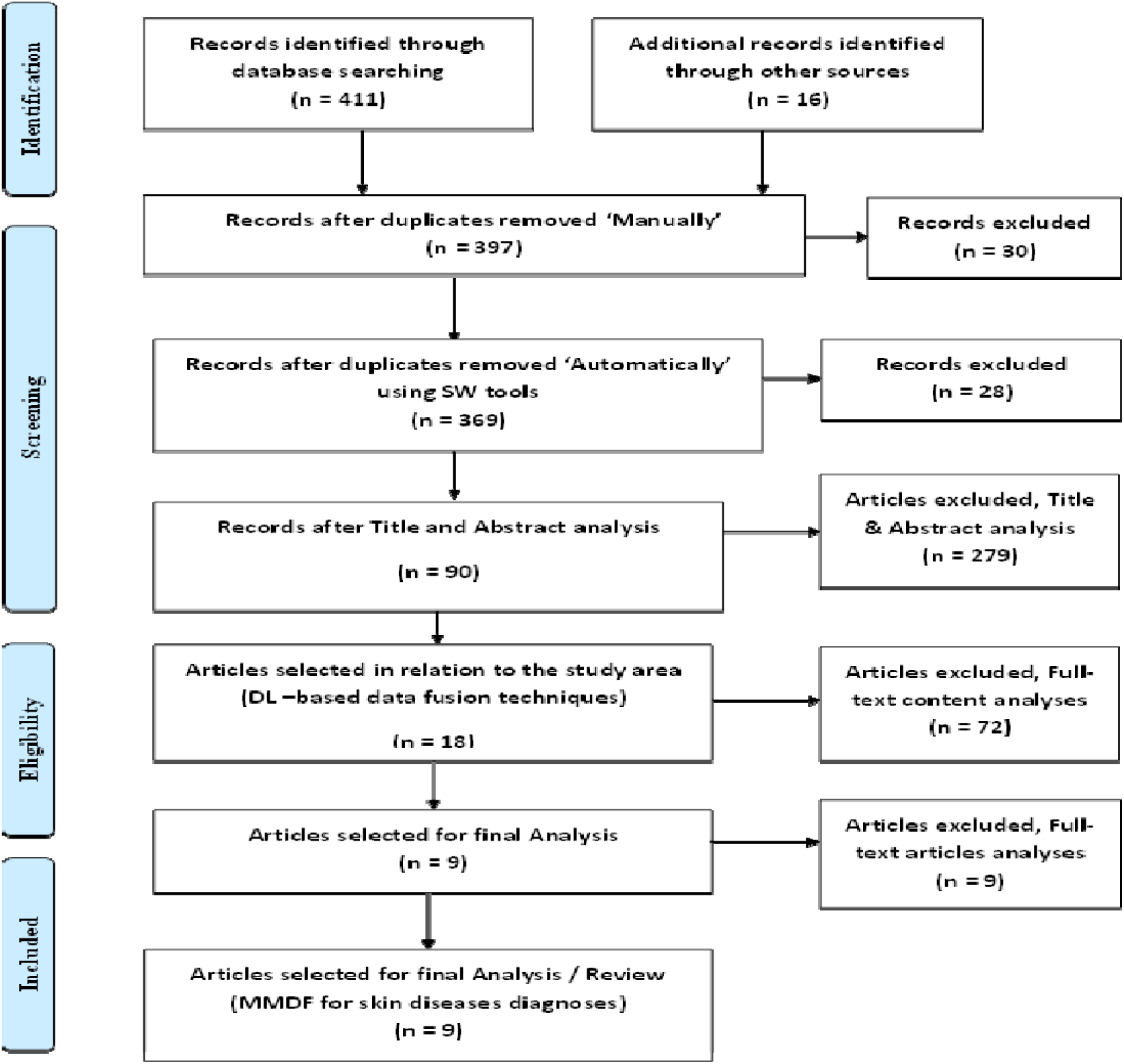
Article selection for systematic literature review following PRISMA flow chart.

## 4. Results

After conducting three levels of screening, 90 articles that have a direct relationship with the current systematic review have been selected for further screening based on full-text reading and analyses. The selected articles and their respective publication year along with the distribution of the publications years have been shown in Figure 4 below. As shown in Figure 4, the article used for this systematic review included studies that have been published recently, where the majority of the studies representing 31% are articles published in 2023, 25% were published in 2022, 16% were published in 2021, 14% were published in 2020, and the remaining 14% were articles published from 2014 – 2019.

**Fig 4:**
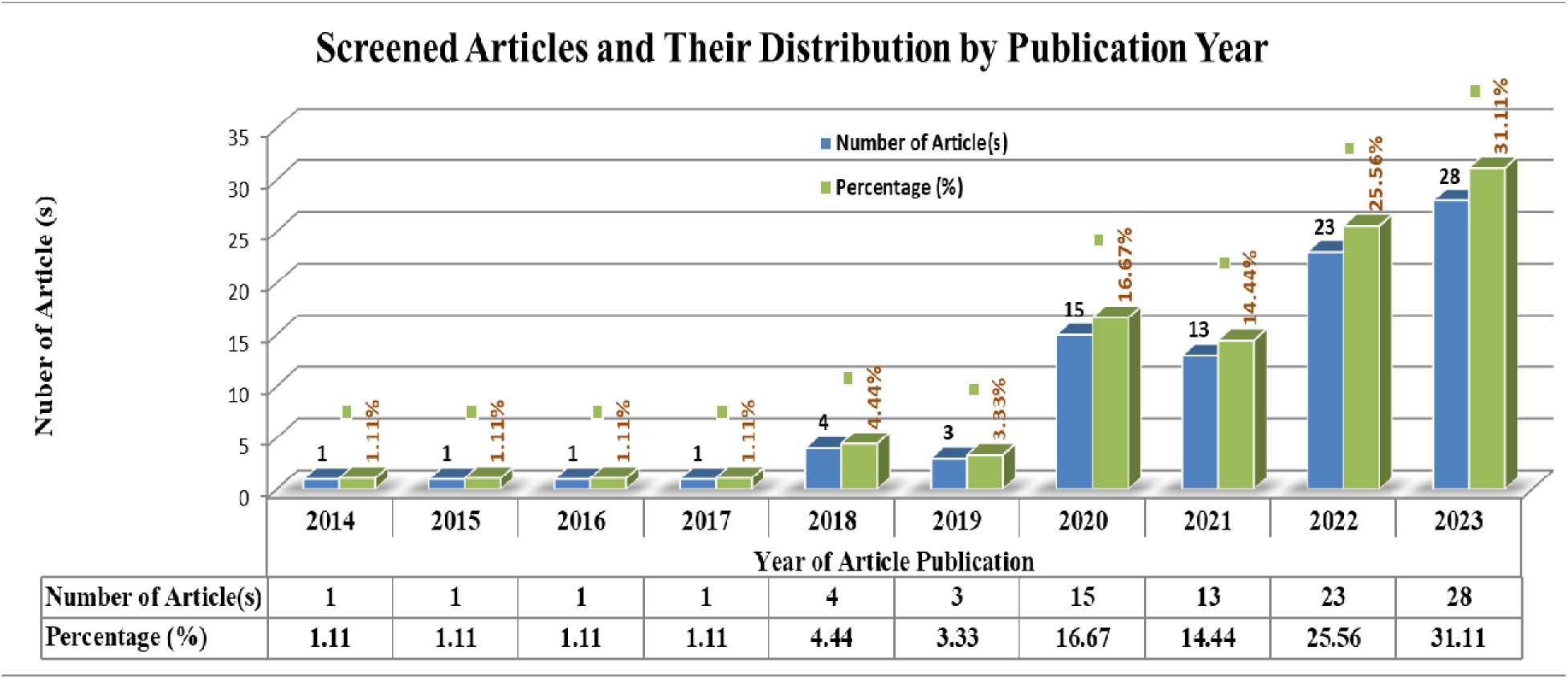
Distribution of articles after the third level screening by publication year.

Finally, the 90 articles were further analyzed by categorizing them into four different groups, (i) articles that utilized DL methods for the diagnosis of skin diseases, (ii) articles that implement ML & DL techniques for the diagnosis of NTDs, (iii) articles about the implementation of multimodal data fusion techniques for medical data fusion, and (iv), articles that implement multimodal data fusion based on DL-based methods for the diagnosis of skin diseases as shown in Figure 5 below.

**Fig 5:**
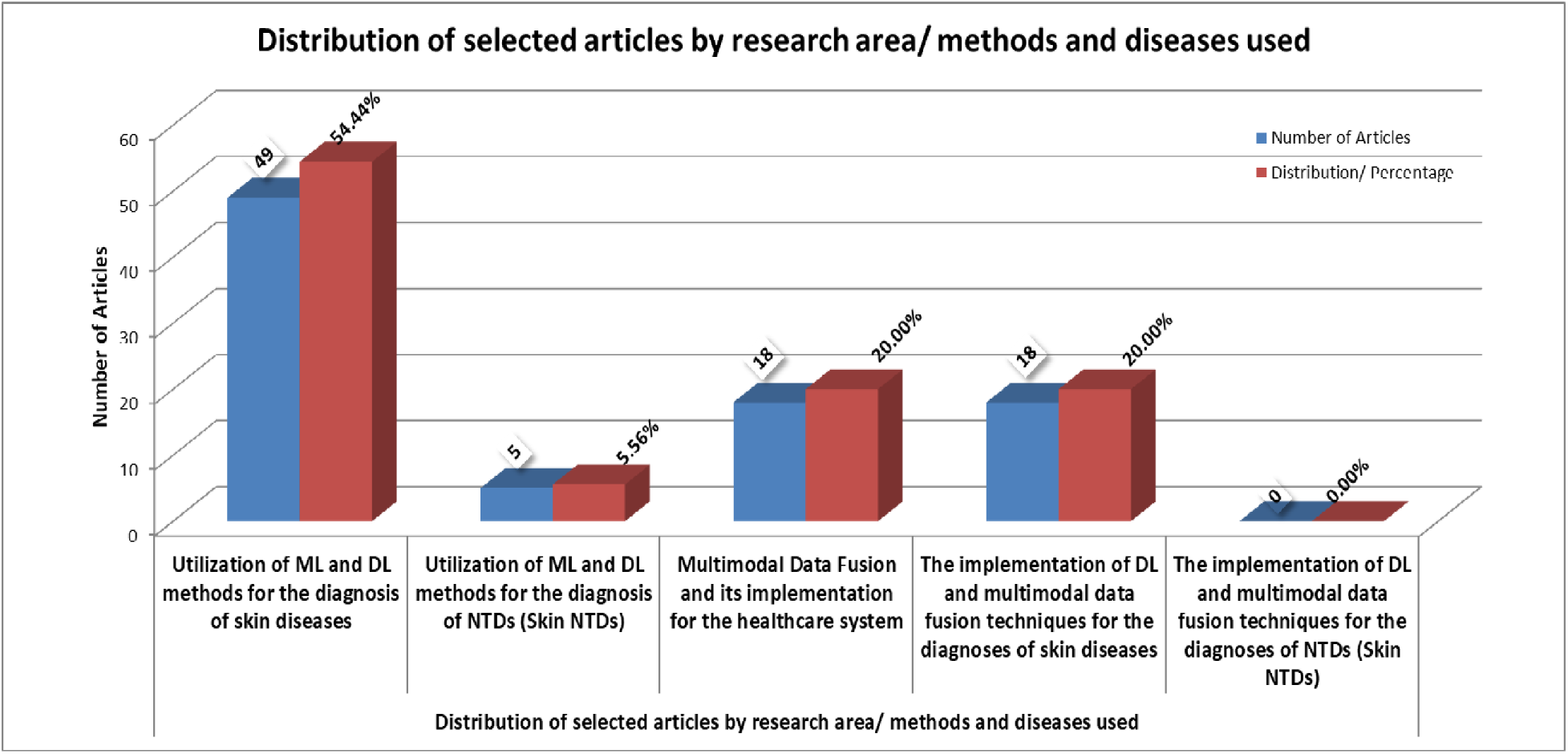
Distribution of articles by research area/methods and selected diseases after the third level screening.

As portrayed in Figure 5 above, 54.44% of articles utilized ML and DL methods for the diagnosis of skin diseases in general, 20% deal with multimodal data fusion techniques for healthcare systems and 20% implementation of DL-based multimodal data fusion methods for the diagnosis of skin diseases. On the other hand, 5.56% of the articles utilized ML and DL methods for the diagnoses of NTDs in general have been identified and analyzed. However, no article has been found that deals with the implementation of DL-based MMDF methods for the diagnosis of NTDs which has led to the analyses of previous studies that used this approach for the diagnoses of different skin diseases other than the NTDs. By conducting the fourth level screening, 18 articles that utilize different fusion techniques for the diagnosis of various skin diseases have been identified.

### 4.1. Article Analysis

The final screening has resulted in the separation of 7 of the 18 articles due to the fusion techniques they utilize for the diagnosis of skin diseases. The fusion techniques presented in those 7 studies are feature fusion (5 studies), image fusion (1 study) and model fusion (1 review study) as presented in Table 1 below. Table 1 presented the analysis of three different types of fusion other than MMDF using five different parameters as shown in the table below.

On the other hand, 2 articles presented a review of the multimodal data fusion techniques for the diagnoses of skin diseases other than NTDs. Although the 2 articles [12][13], didn’t implement MMDF techniques for a specific skin disease diagnosis using their datasets of preferences, they presented theoretical analyses. All in all, 9 articles are used for the final analysis of this review.

After conducting the final screening procedures, 9 articles have been selected for the final analysis of this systematic review as presented in Table 2 below. The 9 articles selected utilized DL-based methods based on MMDF techniques for the diagnoses of different skin diseases other than NTDs. The 9 studies are selected for the final analysis of this review since there are no similar studies found for the diagnosis of skin related NTDs based on MMDF. Since skin related NTDs are being diagnosed using skin photos or images, patient records and related information, these studies are selected and reviewed to analyze the different techniques utilized by those studies. The final analysis is conducted on the 9 articles using 5 different analysis criteria (the methods used, diseases selected for diagnosis, dataset used, algorithms used and corresponding performance achievements) to identify research gaps as presented in Table 2 below.

## 5. Discussion

The primary goal of this systematic review was to collect and analyze research studies that are pertinent to the area of DL-based models that use multimodal data fusion techniques for the diagnosis of skin related NTDs. Since no studies were found in the specified area, similar or related studies that implement MMDF techniques based on DL for the diagnoses of skin diseases were collected and analyzed to extract pertinent information. In doing so, 9 articles (indexed in Scopus and Web of Science) about data fusion techniques, particularly MMDF techniques for the diagnosis of skin diseases have been examined and analyzed. Table 2 below presents the summary of the studies that implemented the different MMDF techniques based on DL for the diagnoses of different skin diseases.

### 5.1. Methods used for building diagnostic models for skin diseases

In the final analysis of this systematic review, the nine studies identified proposed and demonstrated the MMDF approach for the diagnosis of different skin diseases using their corresponding datasets. The studies utilized different methods and algorithms that include CNN, random forest, multilayer perceptron (MLP), long-short term memory (LSTM), the color constancy algorithm, and hyperparameter optimization (HPO) algorithms. Accordingly, 88.9% of the studies (8 articles) primarily utilized the CNN algorithm along with CNN architectures, while 11.1% of the studies utilized MLP and LSTM along with CNN architectures including ResNet50, VGG16, and AlexNet. In general, the studies employed different methods to demonstrate the DL-based methods for combining different modalities of patient data using different methods, such as the attention-based mechanism for combining images and metadata features, a multimodal transformer using the Vision Transformer (ViT) model, and mapping heterogeneous data features. In addition, DCNN architectures such as Densenet121, ILSVRC 2015, VGG16, VGGNet19, ResNet50, ResNet101, DenseNet121, Inception-V3, AlexNet with MLP, AlexNet with LSTM, ResNet50 with MLP, and ViT models were utilized for feature extraction and transfer learning purposes.

### 5.2. Fusion strategies suggested for skin disease diagnosis

Generally, data fusion techniques determine some issues, including the method of integrating data, the data being fused or integrated, and the level at which data will be integrated. The studies used for this review demonstrated various fusion approaches, mainly feature fusion, model fusion, image fusion, and MMDF techniques. In this regard, 89% of the selected studies analyzed in this review implemented MMDF approaches for integrating mainly clinical images and textual medical data. Whereas only one study (11%) demonstrated the MMDF approach for combining two imaging modalities (dermatoscopic and macroscopic images) with patient metadata [35].

As reported by the studies used in this review, various fusion strategies have been experimented with on a particular dataset while developing a diagnostic model for specific skin disease(s). Accordingly, the fusion methods or strategies include integrating multiple imaging modalities (2 image modalities in this case) with textual patient data [35], using a multiplication-based fusion approach (used to control data imbalance) [36], using the metadata processing block (MetaBlock) for enhancing features extracted from the images throughout the classification [7], other study used a naive combination of the patient data classifier module and a whole slide image classifier module [8]. Furthermore, using a DNN that has two encoders for extracting image features and textual features, a MMDF module with intra-modality self-attention and inter-modality cross-attention capability was experimented with, and it was reported that the model outperformed other fusion models [37]. On the other hand, a neural network with a multimodal transformer consisting of two encoders for both images and metadata and one decoder to fuse the multimodal information using the ViT model to extract image features, a soft label encoder for the metadata, and a mutual attention block to fuse the different features [38]. In another study, a fusion system was developed using four procedures consisting of preprocessing the image and metadata, feature extraction using six pre-trained models, feature concatenation (using CNN through convolutional, pooling, and auxiliary layers), and finally classification of skin disease [39]. Similarly, the feature concatenation method was used to develop a wound classifier multimodal network by concatenating the image classifier and location-based classifier outputs [9]. Finally, a skin cancer diagnostic model was developed following three procedures, including extracting features (skin images and patient clinical data using CNN architectures), using the attention mechanism (for handling the multimodal features), and finally developing a feature fusion model [40].

### 5.3. Achievements of MMDF techniques in diagnosing skin diseases

As stated by the studies reviewed, in developing diagnostic models using MMDF techniques for skin diseases, various DL methods and algorithms were used, including CNN, Random Forest, MLP, and LSTM. The algorithms achieved sufficiently higher performances in their respective studies while being tested on a particular dataset. Consequently, it was confirmed that MMDF techniques outperform traditional baseline diagnostic approaches [7][36]. Furthermore, the majority of the studies reviewed reported that the disease classification models achieved accuracy of more than 80% [8][9][35][38]. A study using a DNN with two encoders that implement a multimodal fusion module with intra-modality self-attention and inter-modality cross-attention reported an accuracy of 76.8% [37]. Similarly, another study used in this review that used medical image analysis based on feature extraction, feature concatenation, and classification or diagnosis methods reported 99.94% accuracy in the classification or diagnosis of seven selected skin diseases. In general, as the analysis results show, MMDF techniques are significantly improving classification accuracies. Therefore, the utilization of multimodal data fusion techniques based on the deep learning methods, algorithms, and models in different settings (such as an ensemble of two or more of those methods, algorithms, and models) is a potential research area that needs further investigation, especially for the diagnosis of NTDs.

## 6. Conclusion

In this systematic review, articles were collected from seven major and reputed sources where 427 study papers were organized, classified, screened and selected to analyze the application of DL-based diagnostic models using multimodal data fusion techniques for the diagnoses of skin related NTDs. Although there are studies that demonstrate the utilization of DL methods for the diagnoses of NTDs, no previous studies were found regarding the implementation of MMDF methods for the diagnoses of NTDs. Similar studies using MMDF for the diagnoses of other skin diseases, such as skin cancer, are reviewed to extract information about the implementation of these methods. In doing so, the selected studies are analyzed using parameters such as research approaches used, disease(s) selected for the study, the dataset used, algorithms used, the performance achieved, and future directions suggested by the study. Accordingly, although all the reviewed studies used diverse research methods and datasets based on their problem, DL-based CNN algorithms were found to be by far the most frequently used algorithm by all studies reviewed. In addition, DNN-based network architectures were widely utilized. In general, the implementation of MMDF methods for the diagnosis of skin diseases significantly enhances the diagnostic performances of models as per different studies reviewed, as confirmed in this review. Hence, utilizing MMDF methods for the diagnoses of skin diseases, particularly for skin related NTDs, would be paramount towards developing DL-based diagnostic models for NTDs.

## Data Availability

No data was collected

## Notes

### Competing Interest Statement

The authors have declared no competing interest.

### Funding Statement

The author(s) received no specific funding for this work.

### Summary of Updates

This version of the manuscript was made to modify the public usage of the manuscript in that any usage of the manuscript, including redistribution, remixing, and other forms of using the document, is prohibited unless using official consent from the author(s).

